# Effectiveness of COVID-19 vaccine booster doses in adults aged 50 years and over during the Omicron period in Victoria, Australia

**DOI:** 10.1101/2025.03.31.25324986

**Authors:** Joshua Szanyi, Yue Yang, Jiaxu Zeng, Chris Clarke, Amanda K Buttery, Tony Blakely

## Abstract

**Background:** Country-specific estimates of COVID-19 vaccine effectiveness (VE) are important for policy making, but analyses of COVID-19 VE in Australia have been limited to date.

**Methods:** We used a modified Cox regression model to estimate the adjusted relative VE of three vs. two and four vs. three COVID-19 vaccine doses against hospitalisation and death due to COVID-19 among Victorians aged ≥50 years after the emergence of the Omicron SARS-CoV-2 variant through the linkage of national and state-wide health and administrative datasets. Analyses were conducted in two periods – 1 December 2021 to 19 June 2022 (Omicron BA.1/2 period; analyses of three vs. two doses), and 20 June 2022 to 7 November 2022 (Omicron BA.4/5 period; analyses of four vs. three doses).

**Results:** Approximately 1.8 million people were included in analyses of three vs. two doses and approximately 1.2 million people were included in analyses of four vs. three doses. Adjusted relative VE against death soon after boosting with a third dose (compared to two doses) in individuals aged ≥65 years in the Omicron BA.1/2-dominant period reached 81.2% (95% confidence interval 76.9–84.6%). There was also evidence for a relative benefit of a third dose in the Omicron BA.1/2 period against hospitalisation and for a fourth dose in the Omicron BA.4/5 period against hospitalisation and death in this age group. In contrast, estimates of relative VE in the 50–64-year age group were highly imprecise. *Conclusions:* These results confirm the benefits of vaccine boosters in the Omicron era for those aged ≥65 years, with the most notable gains evident from a third dose in late 2021 to mid-2022.

## Introduction

In May 2023, the World Health Organization announced that COVID-19 was no longer a public health emergency of international concern, signalling a transition to managing COVID-19 alongside other communicable diseases.^1^ A similar sentiment was reflected in Australia’s 2023 National COVID-19 Management Plan,^2^ which emphasised ongoing vaccination as a cornerstone of the long-term COVID-19 response.^1,2^

To inform evidence-based vaccine policy decisions moving forward, policy makers require access to accurate and locally contextualised data regarding COVID-19 vaccine effectiveness (VE). However, there is a relative paucity of Australian VE data available in the published literature, and Australian studies published to date mainly focus on narrow population groups (e.g., people aged 65 years and over) or broad clinical outcomes (e.g., composite outcomes of hospitalisation and death).^3-6^ Accordingly, Australian COVID-19 vaccination policy largely draws on international studies that may not fully reflect the local setting.

There is therefore a need to develop local capability to estimate COVID-19 VE in Australia, but this is a gap in Australia’s current pandemic preparedness and response capacity.^7^ Addressing this gap would benefit not only the COVID-19 response but could also be leveraged to evaluate the effectiveness of vaccines for other respiratory pathogens such as influenza and respiratory syncytial virus and build analytical infrastructure that could be deployed more rapidly in a future pandemic.^8^ In addition, prior to the emergence of the Omicron variant the majority of the Australian population was SARS-CoV-2 infection-naïve,^3^ which provides an opportunity to develop novel insights into COVID-19 VE through the analysis of Australian data.

The aims of the current study were therefore to estimate the relative VE and waning of three vs. two and four vs. three doses of COVID-19 vaccine against hospitalisation and death due to COVID-19 among Victorians aged 50 years and older after the emergence of the Omicron SARS-CoV-2 variant, through the novel linkage of national and state-wide health and administrative datasets.

## Methods

### Study setting

This study was conducted in Victoria, Australia, which has a population of 6.96 million people.^9^ In 2020 and 2021, prior to the study period, there was minimal SARS-CoV-2 transmission in Australia compared to most other countries. Coinciding with the lifting of most travel restrictions and other public health measures, the emergence of the Omicron SARS-CoV-2 variant in late 2021 was followed by extensive SARS-CoV-2 transmission in Victoria such that serological evidence of prior infection in the state rose to approximately 70% of individuals by late 2022.^10^

Australia’s COVID-19 vaccine rollout commenced in February 2021 with Comirnaty (BNT162b2; Pfizer/BioNTech) recommended for all eligible people aged ≥16 years, followed by Vaxzevria (ChAdOx1; AstraZeneca) recommended for all eligible people aged ≥18 years in March 2021. In September 2021, Spikevax (mRNA-1273; Moderna) was recommended for all eligible people aged 12 years and over.^11^ From late 2021 all adults were recommended to have a booster vaccine dose (for most people, a third dose) if they had completed their primary course of vaccination at least 6 months prior. In March 2022, an additional ‘winter booster dose’ (a fourth dose for most people), was recommended for some groups, including people ≥65 years of age, residents of aged/disability care facilities, severely immunocompromised individuals ≥16 years, and Aboriginal and Torres Strait Islander people ≥50 years of age.^12^ In July 2022 all adults ≥30 years of age became eligible for a winter dose.^13^ As of October 2024, Australians aged 75 and older are recommended to have a COVID-19 vaccine booster every six months. Those aged 65 to 74 years, and individuals 18 to 64 who are severely immunocompromised, are recommended to receive a booster annually (but are eligible for a booster every six months).^14^

At the beginning of the study period (1 December 2021) the SARS-CoV-2 Delta variant was detected in almost all sequenced samples in Australia. In early January 2022, the Omicron BA.1 variant became dominant, followed by BA.2 in March 2022. The BA.4 and BA.5 variants became dominant in June 2022 with the BA.5 variant detected in most sequenced samples in Australia until November 2022.^15^ Therefore, two distinct study periods were defined for this study: 1 December 2021 to 19 June 2022 (Omicron BA.1/2 dominant period) to investigate relative VE of three vs. two doses and 20 June 2022 to 7 November 2022 (Omicron BA.4/5 dominant period) to investigate relative VE of four vs. three doses.

### Study design and data sources

This analysis drew on multiple national and Victorian data sources which are fully described in the Supporting Information. Briefly, these included COVID-19 vaccination data from the Australian Immunisation Register (AIR), hospital admission data (from the Victorian Admitted Episodes Dataset), emergency department data (from the Victorian Emergency Minimum Dataset), mortality data (from the Victorian Deaths Index) and COVID-19 testing and other surveillance data (from the Transmission Response Epidemiology Victoria database – a population-based surveillance system capturing COVID-19 data reported by medical practitioners and pathology services in Victoria). Additionally, the Victorian Linkage Map was used to define Victorian residence during the study period. Data linkage was performed by the Centre for Victorian Data Linkage at the Victorian Department of Health.

A cohort of individuals aged ≥50 years (on 1 December 2021) was developed by including those with a record of at least two doses of a COVID-19 vaccination by the end of the study period (7 November 2022) and at least one of:

- any activity recorded within the Victorian Linkage Map during the three years before 1 December 2021 (1 December 2018 to 30 November 2021 inclusive); or
- a record for a non-COVID-19 vaccine during the same three-year period where all recorded places of residence for these vaccination encounters were within Victoria.

Individuals for whom less than 28 days had elapsed between doses two and three or doses three and four were excluded due potential data entry or linkage errors.

Exposure was defined as the receipt of a monovalent COVID-19 vaccination booster. Those who had received three doses were compared to those who had received two doses and those who has received four doses were compared to those who had received three doses. The two clinical outcomes investigated were hospitalisation due to COVID-19 (hospital admission with COVID-19 as a principal or other diagnosis as indicated by International Classification of Diseases 10^th^ Revision codes U07.1, U07.2, U07.11, or U07.12)^16^ and death due to COVID-19 using the Victorian Department of Health’s surveillance definition of a reportable COVID-19 death (see Supporting Information).

### Statistical analyses

A modified Cox regression model,^17-19^ allowing for crossover of participants from the control to the exposed group (for example, from dose two to three), was used to determine the relative VE associated with receipt of a COVID-19 vaccine booster dose. This method has been used previously to investigate the durability of vaccine protection against Omicron subvariants.^20^ Analyses were performed separately for two age groups (≥65 years and 50 to 64 years). For the primary analyses, VE was defined as one minus the hazard ratio, which gives the percentage reduction in the instantaneous risk of disease at day *t* for those who received a booster *t* days ago compared with those who had not received a booster dose.

VE estimates and corresponding 95% confidence intervals (CIs) were modelled as piecewise linear functions of time since each vaccination, with change points placed at 14 and 28 days since boosting. Separate analyses were conducted in the two time periods, 1 December 2021 to 19 June 2022 (Omicron BA.1/2 period; 200 days), and 20 June 2022 to 7 November 2022 (Omicron BA.4/5 period; 140 days), with observational calendar time starting on the first day of the study period and vaccinated time representing the time (in days) since the receipt of a given vaccination dose (dose three or four). Individuals receiving booster doses outside of the two specified time periods were not included in the analysis.

We adjusted for the following covariates: age in years (as a numeric variable), gender, socio-economic status (index of relative socio-economic disadvantage quintile,^21^ as a categorical variable), Aboriginal and/or Torres Strait Islander identity, whether the individual’s primary vaccination course was with an mRNA or non-mRNA vaccine (for comparisons of dose two vs. three only; primary vaccine courses including one dose of an mRNA vaccine and one dose of a non-mRNA vaccine were classified as mRNA primary vaccine courses for the purposes of this analysis), and Charlson comorbidity index as recorded in hospital admissions data from January 2007 onwards (dichotomised as 0 or ≥1, with individuals without a recorded Charlson comorbidity index assigned a value of 0).

Individuals were censored at death due to any cause, receipt of a subsequent booster dose (for example, receipt of a fourth vaccine dose in the analysis of three vs. two vaccine doses), receipt of a bivalent COVID-19 booster dose, or the study end date (7 November 2022), whichever came first. For the hospitalisation outcome, death is a competing risk event. Accordingly, in analyses of VE against hospitalisation we report the number of participants censored due to death from any cause.

All regression analyses were performed in R (version 4.3.0) using the *dove2* command in the *Durability of Vaccine Efficacy (DOVE)* package.^17-19,22^

### Sensitivity analyses

As a sensitivity analysis, VE was alternatively defined as one minus the attack rate, which gives the percentage reduction in the cumulative incidence of disease over *t* days for those who received the third or fourth vaccination *t* days ago compared with those who remain double or triple vaccinated respectively.^19^ We also repeated the primary analyses for individuals aged ≥65 years stratified by the brand of booster (Comirnaty or Spikevax), and with additional change points at four-weekly intervals to test our assumption that VE monotonically decreases with a constant slope on the logarithmic scale after 28 days.

In our primary analyses there was potential for variation in time intervals from receipt of the last vaccine dose between the boosted and comparison groups. Therefore, we also conducted a sensitivity analysis among 65-to 84-year-olds for three vs. two vaccine doses with event counts and person time cross-classified by calendar month, month since receipt of the booster dose among boosted individuals, and month since last dose in the comparator population. We restricted comparator person time to various time lags in their receipt of the comparator dose to the date of receipt of the booster dose among the boosted population (for example, four to seven months prior to receipt of the booster dose). This allowed estimation of the effect of three vs. two doses of vaccine for the three-dose group compared to people who would have been eligible for a third dose at approximately the same time that the three-dose group received their third dose. Finally, we also restricted the third dose group to have had their second dose in the same window as the comparator second dose group, to formally adjust for confounding by time since second dose.

### Ethics approval

Ethics approval for the study was granted by the University of Melbourne Human Research Ethics Committee (2023-24212-40704-5). Custodians responsible for each data set used in the study also provided permission for their use.

## Results

The cohort meeting the study inclusion criteria on 1 December 2021 comprised 1,895,148 individuals (Figure 1).

**Figure 1.**
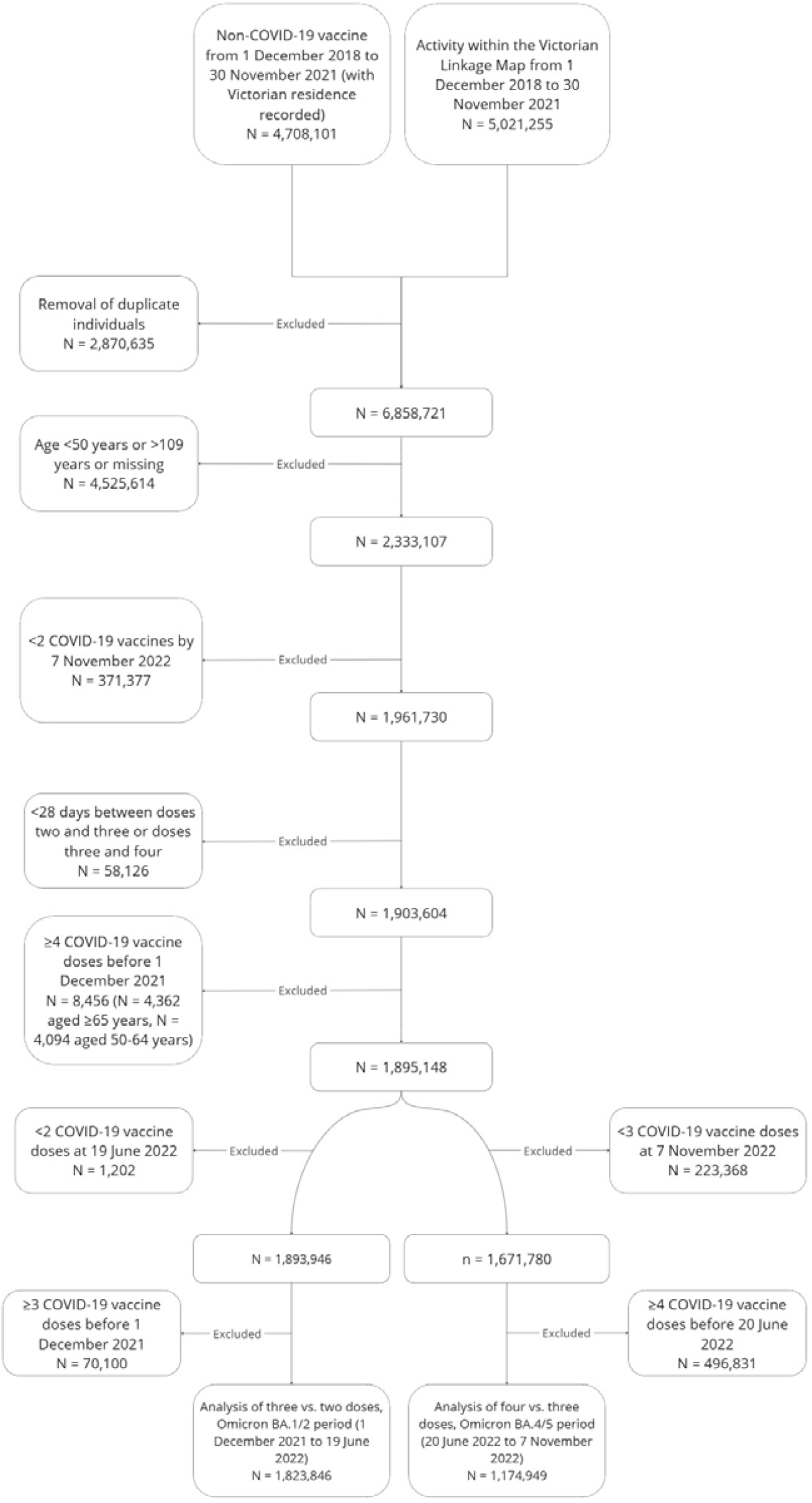
Study cohort selection diagram.

Of these eligible individuals, 1,823,846 were included in the analysis of three vs. two vaccine doses between 1 December 2021 and 19 June 2022, and 1,174,949 were included in the analysis of four vs. three vaccine doses between 20 June and 7 November 2022. The characteristics of the participants included in each of these analyses are summarised in Table 1.

**Table 1:** Cohort characteristics when entering each follow-up period.

|  | Omicron BA.1/2 period (1 December 2021 to 19 June 2022),<br>N = 1,823,846 | Omicron BA.4/5 period (20 June 2022 to 7 November 2022), N =<br>1,174,949 |
| --- | --- | --- |
| *Age* |  |  |
| 50–64 | 868,067 (47.6%) | 711,975 (60.6%) |
| ≥65 | 955,779 (52.4%) | 462,974 (39.4%) |
| *Gender* |  |  |
| Female | 966,312 (53.0%) | 630,661 (53.7%) |
| Male | 857,439 (47.0%) | 544,226 (46.3%) |
| Missing | 95 (0.0%) | 62 (0.0%) |
| *Aboriginal and/or Torres Strait Islander identity* |  |  |
| Not Aboriginal or Torres Strait Islander | 1,798,424 (98.6%) | 1,159,174 (98.7%) |
| Aboriginal and/or Torres Strait Islander | 24,794 (1.4%) | 15,454 (1.3%) |
| Missing | 628 (0.0%) | 321 (0.0%) |
| *Index of relative socio-economic disadvantage quintile* |  |  |
| 1 | 241,016 (13.2%) | 154,628 (13.2%) |
| 2 | 292,753 (16.1%) | 187,958 (16.0%) |
| 3 | 346,435 (19.0%) | 227,085 (19.3%) |
| 4 | 475,438 (26.1%) | 307,663 (26.2%) |
| 5 | 460,373 (25.2%) | 292,512 (24.9%) |
| Missing | 7,831 (0.4%) | 5,103 (0.4%) |
| *Charlson comorbidity index<sup>§</sup>* |  |  |
| 0 | 1,288,814 (70.7%) | 871,544 (74.2%) |
| ≥1 | 535,032 (29.3%) | 303,405 (25.8%) |
| *Primary course included at least one mRNA vaccine* |  |  |
| Yes | 452,977 (24.8%) | 318,436 (27.1%) |
| *Time since baseline dose<sup>#</sup>* |  |  |
| 0–89 days | 3,299 (0.2%) | 97,047 (8.3%) |
| ≥90 days | 1,820,617 (99.8%) | 1,077,902 (91.7%) |
| *COVID-19 vaccination status<sup>†</sup>* |  |  |
| 2 doses | 1,820,616 (99.8%) | 0.00 (0.0%) |
| 3 doses | 3,230 (0.2%) | 1,168,271 (99.4%) |
| 4 doses | 0.0 (0.0%) | 6,678 (0.6%) |

<sup>§</sup>From hospital admissions data (2007 onwards), with missing values reclassified as 0 for all analyses (n = 302,180 [16.6%] for analyses in the Omicron BA.1/2 period, n = 213,589 [18.2%] for analyses in the Omicron BA.4/5 period). <sup>#</sup>For period one this is the time between dose two and entering the window, for period two this is the time between dose three and entering the window. <sup>†</sup>When entering the window.

In analyses of four vs. three vaccine doses there was a predominance of individuals aged 50 to 64 years, and a greater proportion of people had received their last vaccine dose more recently (less than 90 days ago) compared to analyses of three vs. two vaccine doses. For all other characteristics the individuals contributing to the analyses in the two periods were similar. For participants included in analyses of three vs. two vaccine doses in the Omicron BA.1/2 period, 24.8% had received a primary vaccine course containing at least one mRNA vaccine (75.0% of participants received a primary course with two doses of Vaxzevria, 24.6% of participants received a primary course with two mRNA vaccines, and 0.2% of participants received a primary course of one dose of Vaxzevria plus one dose of an mRNA vaccine).

During the study period, there were a total of 27,761 COVID-19 hospitalisations (21,025 in ≥65-year-olds and 6,736 in 50–64-year-olds), and 3,544 COVID-19 deaths (3,344 in ≥65-year-olds and 200 in 50–64-year-olds) in the study cohort. Splitting by analytical period, 17,866 (64.4%) of these hospitalisations and 1,950 (55.0%) of these deaths occurred between 1 December 2021 and 19 June 2022, with the remainder occurring between 20 June 2022 and 7 November 2022 (Figure 2). For analyses of three vs. two vaccine doses against hospitalisation, 1,606 people aged 50–64 years and 13,561 people aged ≥65 years were censored due to death from any cause. During analyses of four vs. three vaccine doses against hospitalisation, 814 people aged 50–64 years and 5,968 people aged ≥65 years were censored due to death from any cause. The median time elapsed between vaccination doses by age and analytical period is shown in Table S1 (Supporting Information).

**Figure 2.**
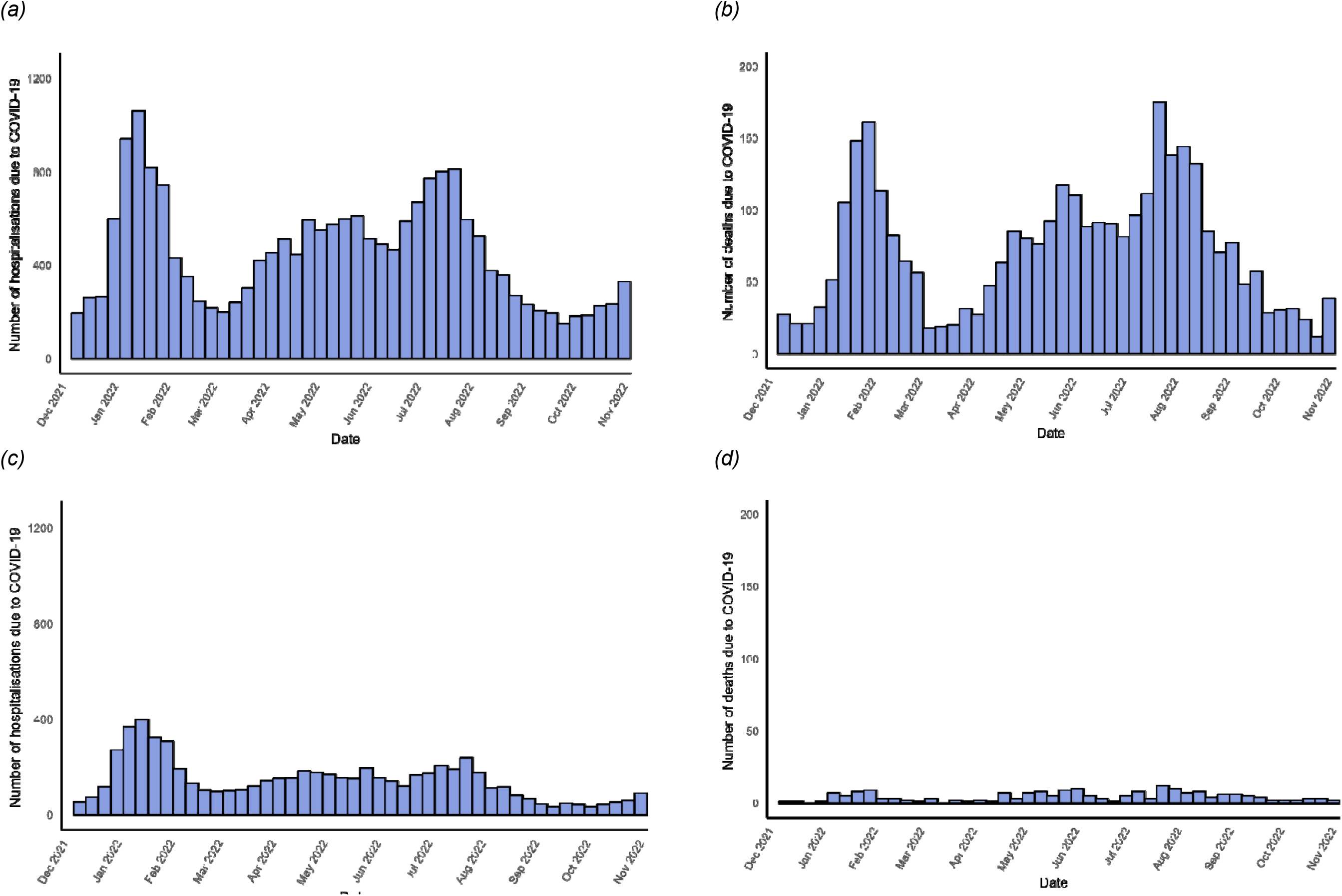
Weekly hospitalisations (a; total = 21,025) and deaths (b; total = 3,344) due to COVID-19 in people aged ≥65 years, and weekly hospitalisations (c; total = 6,736) and deaths (d; total = 200) due to COVID-19 in people aged 50–64 years between 1 December 2021 and 7 November 2022.

Vaccination uptake over the study period is shown in Figure 3. Most people in the 50–64-year age group received their third dose during the first analytical period (1 December 2021 to 19 June 2022) and their fourth dose in the second period (20 June 2022 to 7 November 2022). However, for the ≥65-year age group, many individuals (approximately 450,000) had already received a fourth vaccine dose before the second analytical period and were therefore censored from these analyses. Vaccine uptake by brand is shown in Tables S2 and S3 (Supporting Information).

**Figure 3.**
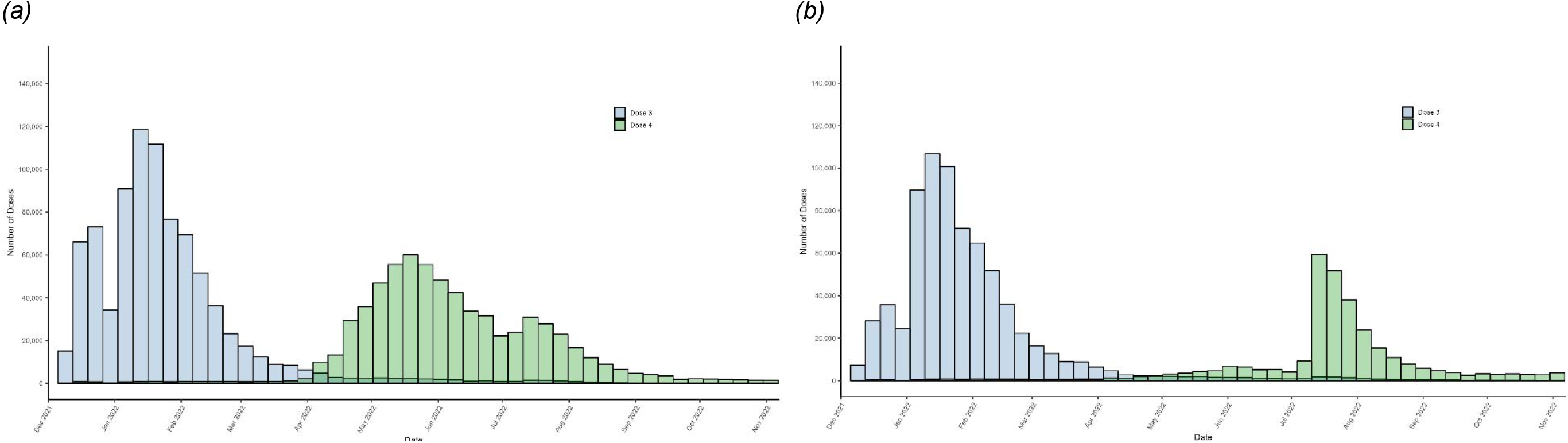
Weekly vaccine uptake between 1 December 2021 and 7 November 2022 in people aged ≥65 years (a) and 50–64 years (b).

Adjusted estimates of relative VE for three vs. two COVID-19 vaccine doses against hospitalisation and death from 1 December 2021 to 19 June 2022 are shown in Figure 4 and Table 2. For the hospitalisation outcome, relative VE of three vs. two doses was highest 28 days following vaccination among individuals aged ≥65 years (63.6%; 95% CI 60.1–66.8%) and 28 days following vaccination among individuals aged 50–64 years (42.0%; 95% CI 32.3–50.3%), falling to zero by 165 and 119 days post-vaccination respectively. For the death outcome, relative VE of three vs. two doses was highest 28 days following vaccination among individuals aged ≥65 years (81.2%; 95% CI 76.9–84.6%) and 14 days following vaccination among individuals aged 50–64 years (64.5%; 95% CI -91.5–93.4%). The former fell to zero by 148 days post-vaccination. Central estimates for those aged 50–64 years remained above zero for the duration of the analysis period, but fewer deaths occurred in this age group and confidence intervals were wide.

**Table 2:** Adjusted relative vaccine effectiveness and 95% confidence intervals from 14 to 180 days post-booster, stratified by outcome, analysis period, booster dose, and age. Models are adjusted for age, gender, socio-economic status, Aboriginal and/or Torres Strait Islander identity, whether the individual’s primary vaccination course was with an mRNA or non-mRNA vaccine (for analyses of dose three vs. two), and Charlson comorbidity index.

| Days | ≥65 years |  |  |  | 50–64 years |  |  |  |
| --- | --- | --- | --- | --- | --- | --- | --- | --- |
|  | Hospitalisation |  | Death |  | Hospitalisation |  | Death |  |
|  | 1 December 2021 to 19 June 2022<br>Three vs. two doses | 20 June 2022 to 7 November 2022<br>Four vs. three doses | 1 December 2021 to 19 June 2022<br>Three vs. two doses | 20 June 2022 to 7 November 2022<br>Four vs. three doses | 1 December 2021 to 19 June 2022<br>Three vs. two doses | 20 June 2022 to 7 November 2022<br>Four vs. three doses | 1 December 2021 to 19 June 2022<br>Three vs. two doses | 20 June 2022 to 7 November 2022<br>Four vs. three doses |
| 14 | 42.9% (34.2–50.4%) | 47.0% (34.3–57.3%) | 79.5% (70.1–85.9%) | 65.5% (42.0–79.5%) | 27.7% (9–42.5%) | 6.2% (-41.9–38%) | 64.5% (-91.5–93.4%) | 89.0% (-222.9–99.6%) |
| 28 | 63.6% (60.1–66.8%) | 48.9% (39.8–56.6%) | 81.2% (76.9–84.6%) | 46.6% (26.3–61.3%) | 42.0% (32.3–50.3%) | 27.8% (-4.1–50%) | 52.4% (-16.6–80.6%) | 35.7% (-130.4–82.1%) |
| 60 | 53.9% (50.5–57%) | 41.3% (34.3–47.5%) | 70.5% (65.6–74.7%) | 40.7% (24.8–53.2%) | 29.6% (20.6–37.6%) | 17.5% (-4.6–34.9%) | 52.8% (11.2–74.9%) | 43.4% (-44.2–77.8%) |
| 90 | 42.5% (38.6–46%) | 33.1% (23.3–41.6%) | 55.1% (48.6–60.8%) | 34.5% (4–55.4%) | 15.7% (4.9–25.2%) | 6.4% (-26.8–31%) | 53.2% (24.1–71.1%) | 49.8% (-149.8–89.9%) |
| 120 | 28.2% (22.7–33.3%) | 23.8% (6.0–38.2%) | 31.6% (20.6–41.1%) | 27.8% (-32.5–60.6%) | -1.1% (-17.6–13.1%) | -6.1% (-72–34.6%) | 53.5% (21.7–72.4%) | 55.5% (-481.9–96.6%) |
| 150 | 10.4% (1.5–18.4%) | N/A | -4.1% (-26.1–14.1%) | N/A | -21.1% (-47.8–0.8%) | N/A | 53.9% (5.3–77.5%) | N/A |
| 180 | -11.9% (-26.2–0.8%) | N/A | -58.4% (-103.0– -23.7%) | N/A | -45.2% (-87.4– -12.5%) | N/A | 54.2% (-22.5–82.9%) | N/A |

**Figure 4.**
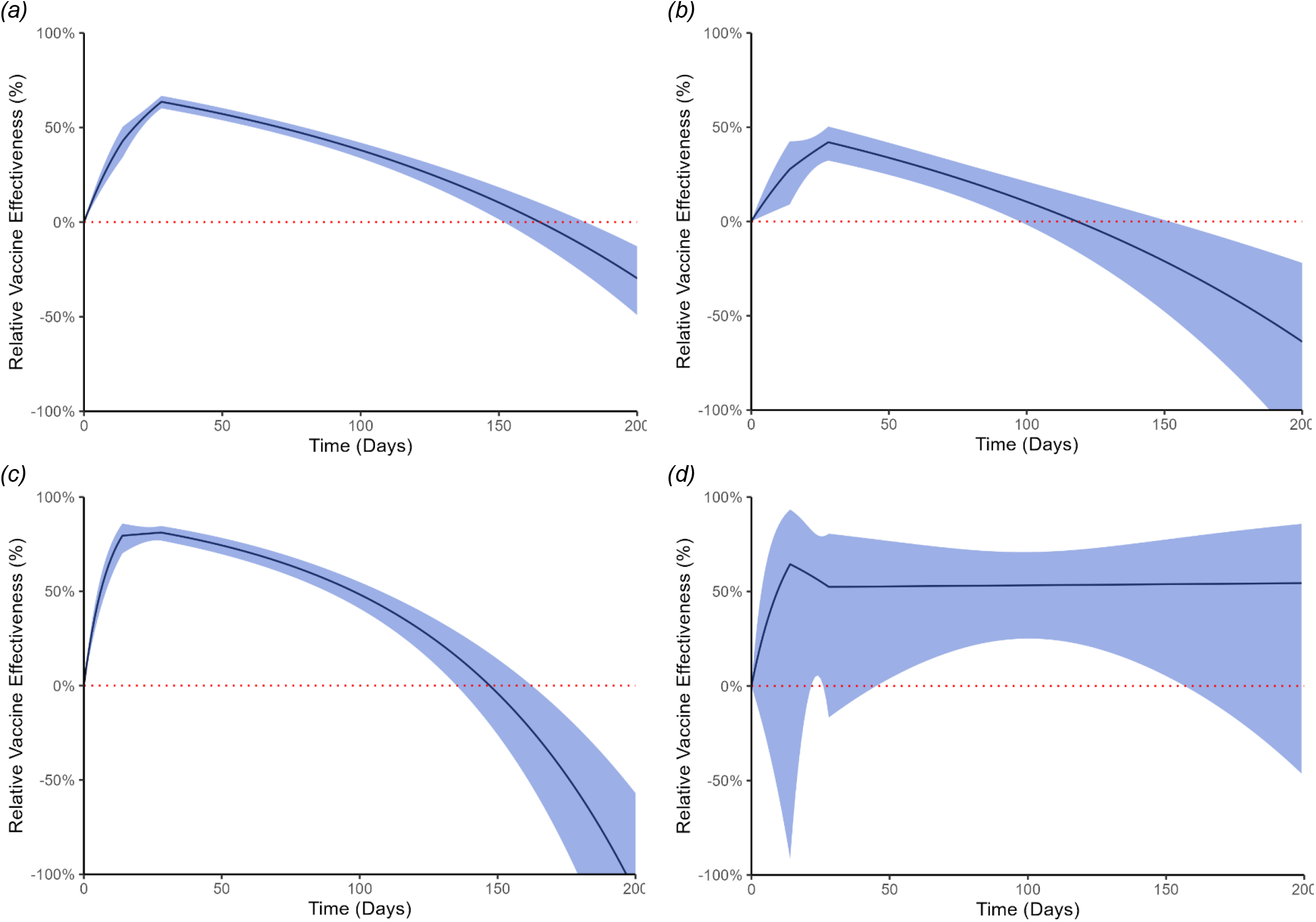
Adjusted relative vaccine effectiveness of three vs. two COVID-19 vaccine doses against hospitalisation for individuals aged ≥65 years (a) and 50–64 years (b), and death for individuals aged ≥65 years (c) and 50–64 years (d) from 1 December 2021 to 19 June 2022. Models are adjusted for age, gender, socio-economic status, Aboriginal and/or Torres Strait Islander identity, whether the individual’s primary vaccination course was with an mRNA or non-mRNA vaccine, and Charlson comorbidity index.

Adjusted estimates of relative VE for four vs. three COVID-19 vaccine doses against hospitalisation and death from 20 June to 7 November 2022 are shown in Figure 5 and Table 2. For the hospitalisation outcome, relative VE of four vs. three doses was highest 28 days following vaccination among individuals aged ≥65 years (48.9%; 95% CI 39.8–56.6%) and 28 days following vaccination among individuals aged 50–64 years (27.8%; 95% CI -4.1–50.0%), although for the latter confidence intervals crossed zero throughout the entire duration of follow-up. For the death outcome, relative VE of four vs. three doses was highest 14 days following vaccination among individuals aged ≥65 years (65.5%; 95% CI 42.0–79.5%) and 14 days following vaccination among individuals aged 50–64 years (89%; 95% CI -222.9–99.6%), but again estimates for the latter were highly uncertain with very wide confidence intervals.

**Figure 5.**
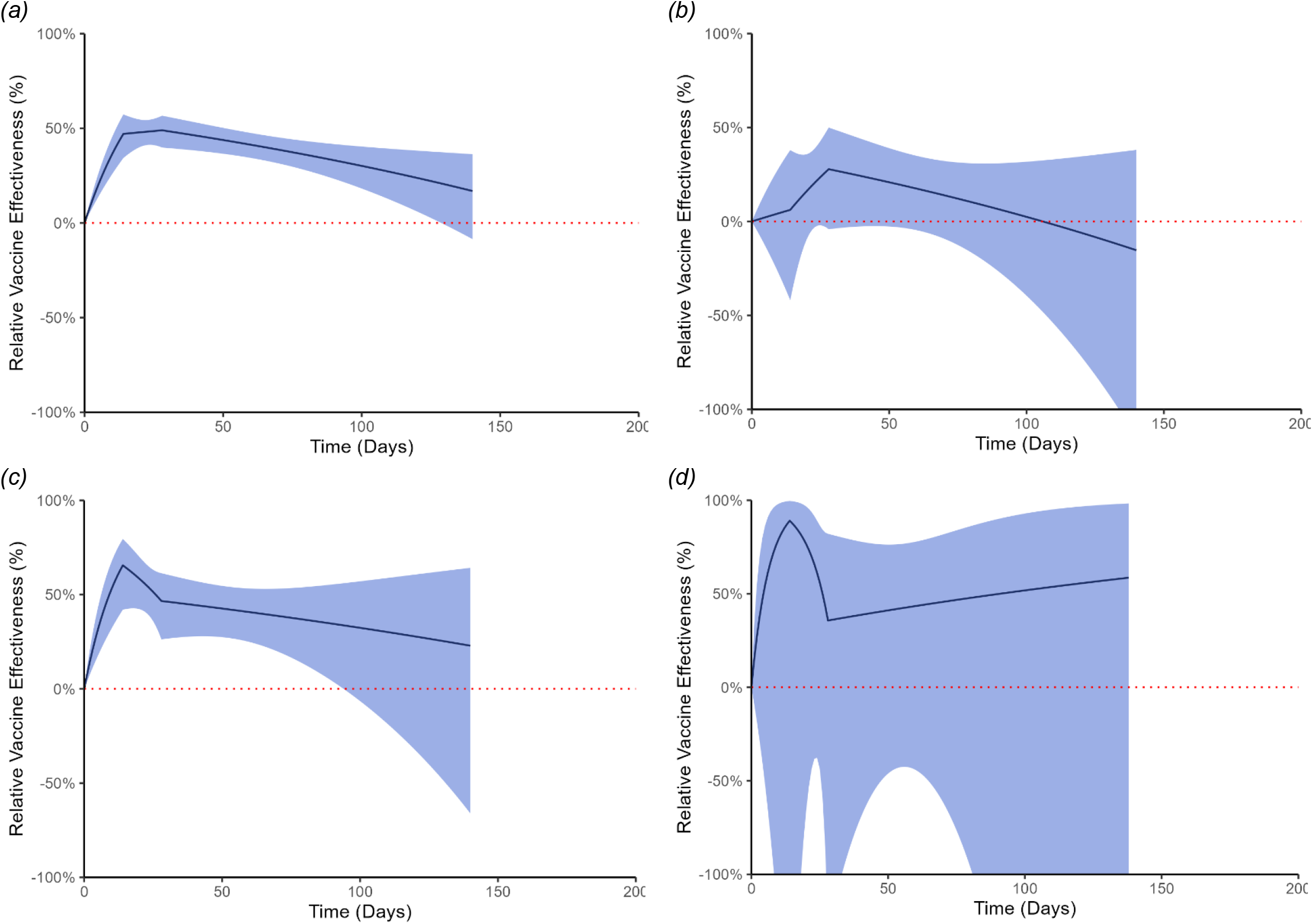
Adjusted relative vaccine effectiveness of four vs. three COVID-19 vaccine doses against hospitalisation for individuals aged ≥65 years (a) and 50–64 years (b), and death for individuals aged ≥65 years (c) and 50–64 years (d) from 30 June 2022 to 7 November 2022. Models are adjusted for age, gender, socio-economic status, Aboriginal and/or Torres Strait Islander identity, and Charlson comorbidity index.

Sensitivity analyses using the attack rate (rather than the hazard ratio) produced similar results to the primary analyses (Supporting Information, Table S4). Similarly, there were negligible differences in VE estimates derived from analyses stratified by time lags between vaccine doses compared to primary analyses (data not shown). In analyses stratified by the brand of booster, VE tended to be higher for Spikevax than Comirnaty, particularly in analyses of three vs. two doses (Supporting Information, Figure S1 and Table S5). For example, relative VE of three vs. two doses against death in this age group peaked at 99.9% (95% CI 94.8–100.0%) 14 days post-booster for Spikevax and 81.3% (95% CI 80.0– 84.8%) 28 days post-booster for Comirnaty. These differences were less pronounced in analyses of four vs. three doses, and there were fewer outcome events observed in this comparative analysis compared to the main analyses (Supporting Information, Tables S6 and S7). When changepoints were placed at four, eight, 12, 16, 20, and 24 weeks, central estimates of relative VE were similar to those in the primary analyses but uncertainty in estimates increased over time since boosting, particularly when VE estimates fell below zero (Supporting Information, Figure S2).

## Discussion

We found that the relative VE of COVID-19 vaccine booster doses varied depending on clinical outcome (hospitalisation or death), age, time since booster dose, and analysis period (Omicron BA.1/2-dominant period or Omicron BA.4/5-dominant period). The highest and most precise level of relative VE was observed against COVID-19 related death soon after boosting with a third dose in those aged ≥65 years in the Omicron BA.1/2-dominant period (peak relative VE >80%), at a time when most older adults received the ancestral strain mRNA vaccine booster in Australia. This relative benefit fell over time since vaccination but persisted for approximately five months compared to receipt of only two doses. Similarly, a fourth ancestral strain monovalent booster dose in the Omicron BA.4/5 period conferred a relative VE of approximately 65% against death in this age group soon after administration compared to a third dose. There was also strong evidence for a benefit of a third and fourth dose against hospitalisation among ≥65-year-olds in the Omicron BA.1/2 and BA.4/5 periods which persisted beyond 160 and 140 days respectively.

In contrast, the relative VE of booster doses in the 50–64-year age group tended to be modest and highly imprecise. However, there were very few outcome events (particularly deaths) which may partially explain the imprecision in VE estimates in this age group. It should be noted that these results may underestimate the benefits of booster doses for immunocompromised people and people with chronic medical conditions that place them at higher risk of severe COVID-19 outcomes, who may have received booster doses earlier than the general population and prior to the follow-up time windows used in this study.

A relatively small number of other studies have evaluated COVID-19 VE in Australia. Our findings are comparable to a study of 2.1 million infection-naïve people aged ≥40 years residing in Sydney, which showed a relative VE for three vs. two doses of 65% (95% CI 61–69%) for a composite outcome of hospitalisation or death soon after vaccination,^3^ and an Australia-wide study using census and other linked data sources reporting relative VE of 64% (95% CI 59–68%) for four vs. three doses against death between June and November 2022 among those aged ≥65 years.^6^ Similarly, a study of 1.1 million people aged ≥65 years in Victoria assessing relative VE between 1 June 2022 to 1 March 2023 reported that a fourth dose reduced hospitalisation or death by 51% (95% CI 42–59%) relative to three doses which waned to 17% (95% CI 3–28%) once ≥25 weeks had elapsed.^23^ Comparisons to international VE studies are challenging due to variations in vaccine policy, circulating variants and methodological differences. However, our results are also similar to those of a meta-analysis of international studies reporting relative VE of three doses vs. primary series of 65.4% (95% CI 53.1–77.6%) against hospitalisation and 76.5% (95% CI 68.2–84.8%) against death.^24^

This study has several strengths, including reporting of VE estimates among people aged 50 years and older, the calculation of VE estimates for discrete rather than composite clinical outcomes (hospitalisation and death), clear delineation of the additional benefits of COVID-19 booster doses by reporting relative rather than absolute VE, and aligning analysis periods with Australian laboratory data on circulating variants. The statistical package used for analysis allowed for the estimation of time-varying VE and accounted for changes in community SARS-CoV-2 transmission during the analysis periods by considering calendar time.

However, this study has several limitations. Firstly, while substantial efforts were made to adjust for residual confounding, limitations in data availability and quality meant that prior SARS-CoV-2 infection, behavioural factors (for example, mask-wearing or healthcare-seeking behaviour), and the presence of pre-existing medical conditions that may influence the timing and likelihood of vaccine uptake and increase COVID-19-related morbidity and mortality were potentially not adequately accounted for. Government recommendations on priority comorbid populations and booster timing intervals changed over time and may have influenced uptake and brand receipt among individuals with comorbidities, warranting cautious interpretation of the results particularly in the stratified analysis by booster brand. Comorbidity is imperfectly captured by the Charlson comorbidity index, which in this instance was calculated only from hospital admissions data and included as a dichotomous variable (as statistical models with more categories generally failed to converge). More comprehensive comorbidity data from, for example, linked primary care records could improve future VE analyses in this context.

Modest selection bias could have been introduced when attempting to restrict analysis to Victorian residents. Because we did not have access to a population spine (based on, for example, census data) that we could link to COVID-19 epidemiologic data, interaction with at least one of the Victorian Linkage Map data sets or receipt of a non-COVID-19 vaccination in the preceding three years were used as proxies for Victorian residence. While the range of data sets in the Victorian Linkage Map is broad (see Supplementary Material), many of these data sets relate to accessing health care services, and it is therefore possible that individuals at risk of more severe clinical outcomes could have been over-represented in the study cohort. Another possible source of selection bias is the fact that individuals were only included in the analysis for a given analytical period if they received their booster during that period (this is particularly relevant for analyses of four vs. three doses for ≥65-year-olds in the Omicron BA.4/BA.5 period, given many people in this age group received a fourth dose prior to 20 June 2022). Furthermore, we cannot rule out potential non-differential misclassification of the hospitalisation or death outcomes (for example, unvaccinated people being more likely to have their hospitalisation attributed to COVID-19 by their treating clinician than vaccinated people).

In our analyses, relative VE reduced over time, eventually falling below zero in some instances (e.g., after 165 days since boosting in the analysis of three vs. two vaccine doses against hospitalisation among people ≥65 years old). Negative VE estimates have been observed in other studies with extended follow-up, and are likely explained by sparse data at longer follow-up times, unmeasured confounding, and bias related to depletion of susceptibles rather than true negative biological effects.^25-27^ Sensitivity analyses with additional changepoints added indicated substantial uncertainty around negative VE estimates (usually with upper confidence limits above zero).

Infection-induced immunity likely increased during the study period due to high levels of SARS-CoV-2 transmission. This could have further lowered VE estimates, because the comparator population may have been more likely than the boosted cohort to have acquired additional immunity from undocumented infection. Those recently infected with SARS-CoV-2 are advised to wait six months before receiving booster doses, also making them more likely to remain in the comparator (that is, less vaccinated) group. We were not able to adequately account for prior infections, which were not routinely reported across the study period unless linked to a hospitalisation or death and are difficult to accurately record without access to longitudinal serological data.

In conclusion, the unique Australian epidemiologic context provides an opportunity to develop novel insights into COVID-19 VE, but such analyses have been relatively limited to date. We used a modified Cox model to determine the relative time-varying VE of COVID-19 booster doses in two distinct periods (an Omicron BA.1/2-dominant period and an Omicron BA.4/5-dominant period) in Victoria and found the most notable benefit of monovalent ancestral booster doses among individuals aged ≥65 years. This analysis was restricted to recipients of ancestral strain-based vaccines, and bivalent and updated variant-focused vaccines may confer greater or more persistent benefit than the original monovalent boosters that were assessed in this study.^28^

This study demonstrates the utility of industry, government and academia collaborating to conduct real-world VE studies, which has been identified as an important current gap in Australia’s pandemic preparedness and response capacity.^7^ To enable timely, evidence-based vaccine recommendations over the coming years, improved access to linked data is needed to facilitate rapid assessment of the relative benefit and optimal timing of updated COVID-19 vaccine boosters designed to more closely match circulating SARS-CoV-2 strains.

## Data Availability

Applications for access to the underlying data used in this study can be made to the Centre for Victorian Data Linkage (https://vahi.vic.gov.au/ourwork/data-linkage). Data cannot be directly shared by the study authors as this is not covered by ethics committee approval or agreements with data custodians.

## Acknowledgements

The authors would like to thank Dr Driss Ait Ouakrim from the University of Melbourne, Dr Suman Majumdar, Daniel West, Sharon Williams, and Caroline Sumpton from the Victorian Department of Health, Dr Andree Hubber, Dr David Martin, Dr Daina Esposito, and Dr Morgan Marks from Moderna Inc, and Dr Katie Mues from Aetion Inc for their valuable inputs into the design and conduct of this study.

