## Supporting information for "Effectiveness of COVID-19 vaccine booster doses in adults aged 50 years and over during the Omicron period in Victoria, Australia"

**Supplementary methods**

Data sources

Data linkage was undertaken by the Centre for Victorian Data Linkage (CVDL) at the Victorian Department of Health. Using the Victorian Linkage Map (VLM), CVDL provided an encrypted identifier for each record in the databases. Access was then provided to the unit level record data within a secure computing environment. Deterministic linkage was undertaken between tables within the supplied database via the encrypted identifier.

*Victorian Linkage Map*

The VLM, a system of linked records across several databases, is maintained by CVDL. In this study, interaction with any of the datasets contained in the VLM during the study eligibility period (three years prior to 1 December 2021) was used as one proxy method for determining current residence in Victoria. These datasets include those related to alcohol and drug services; public and private hospital admissions; births; cancer; family services (including child protection services, family violence, and sexual assault support services); community health services; deaths; disability services; elective surgery waiting lists; emergency department presentations; non-admitted hospital services; home and community care; homelessness services; public housing; mental health services; perinatal services; notifiable infectious diseases; and radiotherapy services. Note no information is received from these datasets (except for those which are used to determine study outcomes) other than an indicator that signifies contact with one or more of the aforementioned services within the eligibility time window.

*Australian Immunisation Register*

The Australian Immunisation Register (AIR) is national register recording vaccines administered to all people in Australia (including those provided under the National Immunisation Program, school programs, or privately). All COVID-19 vaccines are recorded by brand and type (e.g., monovalent, bivalent). For this study, COVID-19 vaccines were identified in the AIR using the following codes: ASTCOV (AstraZeneca Covishield); COVAST (AstraZeneca Vaxzevria); BHACOV (Bharat Biotech Covaxin); GAMSPU (Gamaleya Sputnik V); JANSSE (Janssen-Cilag COVID Vaccine); MODBIV (Moderna Spikevax Biv BA.1); MODBBA (Moderna Spikevax Biv BA.4-5); MODERN (Moderna Spikevax); NOVNUV (Novavax Nuvaxovid); COMBIV (Pfizer Comirnaty Biv BA.1); COMBBA (Pfizer Comirnaty Biv BA.4-5); COMIRN (Pfizer Comirnaty); SINBBI (Sinopharm BBIBP-CorV); SINCOR (Sinovac Coronavac); MODPED.^1^ Note only monovalent vaccines were included in the final analysis, with receipt of a bivalent vaccine (MODBIV, MODBBA, COMBIV, COMBBA) resulting in censoring.

*Victorian Emergency Minimum Dataset*

This dataset includes demographic, administrative and clinical data relating to presentations at Victorian public hospital emergency departments.

*Victorian Admitted Episodes Dataset*

This dataset includes a minimum set of data for each admitted patient episode in all Victorian public and private hospitals.

*Victorian Deaths Index*

This dataset contains data derived from the registry of births deaths and marriages regarding death (including date and cause of death).

*Transmission and Response Epidemiology Victoria*

This is a database containing data related to COVID-19 in Victoria, including SARS-CoV-2 test results and clinical outcomes such as death due to COVID-19.

Outcome definitions

1. Hospitalisation: hospital admission with COVID-19 as a principal or additional diagnosis (ICD-10 codes U07.1, U07.2, U07.11, or U07.12).
2. Death: death where the direct, antecedent or other cause of death is stated as COVID-19 by the notifying doctor, coroner or via Medical Certificate Cause of Death (1a, b, c, d or 2 in coroner report of death); *or* the direct cause of death (on the Medical Certificate Cause of Death) is pneumonia (excluding aspiration pneumonia), bronchopneumonia, pneumonitis, chest infection, lower respiratory tract infection, or respiratory failure and death has occurred within 35 days after diagnosis of COVID-19; *or* the cause of death does not specify COVID-19 on the Medical Certificate Cause of Death but the death occurs up to 35 days after diagnosis of COVID-19 from any cause except respiratory conditions (see above) or incidental conditions such as trauma (excluding falls in the elderly), suicide, motor vehicle accident, or other unrelated accident/injury including drowning, burns, drug, toxin, or poisoning. This definition aligns with the Victorian Department of Health’s surveillance definition of a COVID-19 death.

**Supplementary results**

Supplementary Table 1: Median time (days) between vaccine doses.

|  | *1 December 2021 to 19 June 2022*  *Dose 2 to 3* | *20 June 2022 to 7 November 2022*  *Dose 3 to 4* |
| --- | --- | --- |
| 50–64 years | 151 days | 195 days |
| ≥65 years | 155 days | 175 days |

Supplementary Table 2: Vaccine doses stratified by brand, for participants ≥65 years of age included in each analysis.

|  | *1 December 2021 to 19 June 2022*  *3 vs 2 doses*  *N = 955,779* | | | *20 June 2022 to 7 November 2022*  *4 vs 3 doses*  *N = 462,974* | | |
| --- | --- | --- | --- | --- | --- | --- |
|  | Dose 2 | Dose 3 | Dose 4 | Dose 2 | Dose 3 | Dose 4 |
| Vaxzevria | 848,736 (88.8%) | 11,077 (1.2%) | N/A | 405,812 (87.7%) | 8,086 (1.7%) | 1,243 (0.3%) |
| Comirnaty | 93,979 (9.8%) | 699,795 (73.2%) | N/A | 50,943 (11.0%) | 373,682 (80.7%) | 178,490 (38.6%) |
| Spikevax | 11,758 (1.2%) | 143,333 (15.0%) | N/A | 5,830 (1.3%) | 78,865 (17.0%) | 44,431 (9.6%) |
| Other | 1,306 (0.1%) | 1,777 (0.2%) | N/A | 389 (0.1%) | 2,341 (0.5%) | 4,707 (1.0%) |
| Not received | 0 (0.0%) | 99,797 (10.4%) | N/A | 0 (0.0%) | 0 (0.0%) | 234,103 (50.6%) |

Supplementary Table 3: Vaccine doses stratified by brand, for participants 50–64 years of age included in each analysis.

|  | *1 December 2021 to 19 June 2022*  *3 vs 2 doses*  *N = 868,067* | | | *20 June 2022 to 7 November 2022*  *4 vs 3 doses*  *N = 711,975* | | |
| --- | --- | --- | --- | --- | --- | --- |
|  | Dose 2 | Dose 3 | Dose 4 | Dose 2 | Dose 3 | Dose 4 |
| Vaxzevria | 519,710 (59.9%) | 6,871 (0.8%) | N/A | 450,184 (63.2%) | 6,958 (1.0%) | 702 (0.1%) |
| Comirnaty | 328,082 (37.8%) | 561,610 (64.7%) | N/A | 254,335 (35.7%) | 555,489 (78.0%) | 177,571 (24.9%) |
| Spikevax | 18,640 (2.1%) | 150,793 (17.4%) | N/A | 7,055 (1.0%) | 146,433 (20.6%) | 73,688 (10.3%) |
| Other | 1,635 (0.2%) | 2,156 (0.2%) | N/A | 401 (0.1%) | 3,095 (0.4%) | 11,186 (1.6%) |
| Not received | 0 (0.0%) | 146,637 (16.9%) | N/A | 0 (0.0%) | 0 (0.0%) | 448,828 (63.0%) |

Supplementary Table 4: Adjusted relative vaccine effectiveness and 95% confidence intervals (one minus the attack rate) from 14 to 180 days post-booster, stratified by outcome, analysis period, booster dose, and age. Models are adjusted for age, gender, socio-economic status, Aboriginal and/or Torres Strait Islander identity, whether the individual’s primary vaccination course was with an mRNA or non-mRNA vaccine (for analyses of dose three vs. two), and Charlson comorbidity index.

| Days | ≥65 years | | | | 50–64 years | | | |
| --- | --- | --- | --- | --- | --- | --- | --- | --- |
|  | Hospitalisation | | Death | | Hospitalisation | | Death | |
|  | *1 December 2021 to 19 June 2022*  *Three vs. two doses* | *20 June 2022 to 7 November 2022*  *Four vs. three doses* | *1 December 2021 to 19 June 2022*  *Three vs. two doses* | *20 June 2022 to 7 November 2022*  *Four vs. three doses* | *1 December 2021 to 19 June 2022*  *Three vs. two doses* | *20 June 2022 to 7 November 2022*  *Four vs. three doses* | *1 December 2021 to 19 June 2022*  *Three vs. two doses* | *20 June 2022 to 7 November 2022*  *Four vs. three doses* |
| 0–14 | 24.8% (19.5–29.7%) | 27.4% (19.6–34.4%) | 51.8% (44.2–58.4%) | 40.2% (25.1–52.3%) | 15.5% (5.2–24.7%) | 3.4% (-20–22.2%) | 39.5% (-26.4–71%) | 61.6% (-22–87.9%) |
| 15–28 | 55.4% (51.4–59.1%) | 48.1% (41.0–54.3%) | 80.4% (75.9–84.1%) | 55.4% (40.9–66.4%) | 36.1% (26.6–44.3%) | 18.9% (-4.7–37.2%) | 57.9% (3.3–81.7%) | 66.1% (-22.2–90.6%) |
| 29–60 | 58.6% (55.3–61.8%) | 45% (37.4–51.6%) | 75.9% (71.3–79.7%) | 43.5% (27.9–55.7%) | 35.6% (26.5–43.6%) | 22.4% (-2.4–41.2%) | 52.6% (-0.1–77.6%) | 39.9% (-55.7–76.8%) |
| 61–90 | 48% (44.5–51.3%) | 37% (29.2–43.9%) | 62.8% (57.3–67.7%) | 37.5% (15.8–53.6%) | 22.4% (12.9–30.9%) | 11.7% (-13.4–31.3%) | 53% (19.8–72.4%) | 46.9% (-78.8–84.2%) |
| 91–120 | 35.1% (30.5–39.4%) | 28.2% (14.7–39.6%) | 43.4% (35–50.7%) | 31% (-13.8–58.2%) | 7% (-6.4–18.7%) | -0.1% (-48.6–32.6%) | 53.3% (24.8–71.1%) | 52.9% (-283.3–94.2%) |
| 121–150 | 19.0% (11.9–25.6%) | N/A | 13.8% (-2.2–27.4%) | N/A | -11.5% (-32.9–6.6%) | N/A | 53.7% (14.5–74.9%) | N/A |
| 151–180 | -1.1% (-12.7–9.3%) | N/A | -31.2% (-63.9– -5%) | N/A | -33.6% (-68.0– -6.2%) | N/A | 54% (-8.1–80.5%) | N/A |

Supplementary Table 5: Relative VE for individuals aged ≥65 years stratified by brand of booster. Models are adjusted for age, gender, socio-economic status, Aboriginal and/or Torres Strait Islander identity, whether the individual’s primary vaccination course was with an mRNA or non-mRNA vaccine (for comparisons of three vs. two doses only), and Charlson comorbidity index.

| Days | Comirnaty | | | | Spikevax | | | |
| --- | --- | --- | --- | --- | --- | --- | --- | --- |
|  | Hospitalisation | | Death | | Hospitalisation | | Death | |
|  | *1 December 2021 to 19 June 2022*  *Three vs. two doses* | *20 June 2022 to 7 November 2022*  *Four vs. three doses* | *1 December 2021 to 19 June 2022*  *Three vs. two doses* | *20 June 2022 to 7 November 2022*  *Four vs. three doses* | *1 December 2021 to 19 June 2022*  *Three vs. two doses* | *20 June 2022 to 7 November 2022*  *Four vs. three doses* | *1 December 2021 to 19 June 2022*  *Three vs. two doses* | *20 June 2022 to 7 November 2022*  *Four vs. three doses* |
| 14 | 48.5% (40.0–55.9%) | 51.5% (38.6–61.8%) | 79.4% (69.9–86.0%) | 62.6% (36.4–78.0%) | 89.7% (84.1–93.3%) | 66.9% (42.15%, 81.11%) | 99.9% (94.8–100.0%) | 97.3% (29.6–99.9%) |
| 28 | 66.7% (63.4–69.8%) | 49.8% (40.4–57.8%) | 81.3% (77.0–84.8%) | 45.9% (24.6–61.3%) | 80.9% (76.9–4.1%) | 66.2% (47.54%, 78.23%) | 97.5% (94.6–98.9%) | 69.2% (17.9–88.4%) |
| 60 | 57.2% (54.0–60.2%) | 41.7% (34.3–48.2%) | 70.6% (65.6–74.9%) | 37.9% (20.6–51.4%) | 73.9% (70.3–77.1%) | 55% (41.1%, 65.59%) | 94.7% (90.8–96.9%) | 69.1% (40.3–84.0%) |
| 90 | 45.8% (42.1–49.2%) | 32.8% (22.4–41.8%) | 55.1% (48.7–60.8%) | 29.2% (-4.3–52.0%) | 65.2% (61.5–68.6%) | 41.1% (21.49%, 55.8%) | 89.1% (83.9–92.6%) | 69.0% (1.3–90.3%) |
| 120 | 31.3% (26.0–36.3%) | 22.6% (3.6–37.8%) | 31.5% (20.5–40.9%) | 19.3% (-48.8–56.3%) | 53.5% (47.6–58.8%) | 22.9% (-22.25%, 51.4%) | 77.6% (67.0–84.8%) | 68.9% (-107.8–95.4%) |
| 150 | 13% (4.2–21.0%) | N/A | -4.7% (-26.8–13.6%) | N/A | 37.9% (26.4–47.7%) | N/A | 54% (21.1–73.2%) | N/A |
| 180 | -10.2% (-24.8–2.7%) | N/A | -59.9% (-105.1– -24.7%) | N/A | 17.1% (-4.7–34.4%) | N/A | 5.6% (-101.8–55.9%) | N/A |

Supplementary Table 6: Number of outcome events among boosted and non-boosted individuals in each analysis, by age group.

| Age group | Hospitalisation | | | | Death | | | |
| --- | --- | --- | --- | --- | --- | --- | --- | --- |
|  | *1 December 2021 to 19 June 2022*  *Three vs. two doses* | | *20 June 2022 to 7 November 2022*  *Four vs. three doses* | | *1 December 2021 to 19 June 2022*  *Three vs. two doses* | | *20 June 2022 to 7 November 2022*  *Four vs. three doses* | |
|  | *Boosted participants* | *Non-boosted participants* | *Boosted participants* | *Non-boosted participants* | *Boosted participants* | *Non-boosted participants* | *Boosted participants* | *Non-boosted participants* |
| 50–64 years | 1,479 | 2,083 | 168 | 869 | 45 | 54 | 7 | 52 |
| ≥65 years | 5,051 | 5,204 | 712 | 2,485 | 786 | 850 | 142 | 502 |

Supplementary Table 7: Number of outcome events among boosted and non-boosted individuals in analyses stratified by brand of booster.

| Booster | Hospitalisation | | | | Death | | | |
| --- | --- | --- | --- | --- | --- | --- | --- | --- |
|  | *1 December 2021 to 19 June 2022*  *Three vs. two doses* | | *20 June 2022 to 7 November 2022*  *Four vs. three doses* | | *1 December 2021 to 19 June 2022*  *Three vs. two doses* | | *20 June 2022 to 7 November 2022*  *Four vs. three doses* | |
|  | *Boosted participants* | *Non-boosted participants* | *Boosted participants* | *Non-boosted participants* | *Boosted participants* | *Non-boosted participants* | *Boosted participants* | *Non-boosted participants* |
| Spikevax (AIR code: MODERN) | 655 | 2,955 | 108 | 2,295 | 39 | 839 | 11 | 499 |
| Comirnaty (AIR code: COMIRN) | 4,269 | 4,827 | 592 | 2,429 | 733 | 850 | 129 | 502 |

| *(a)* | *(b)* |
| --- | --- |
| 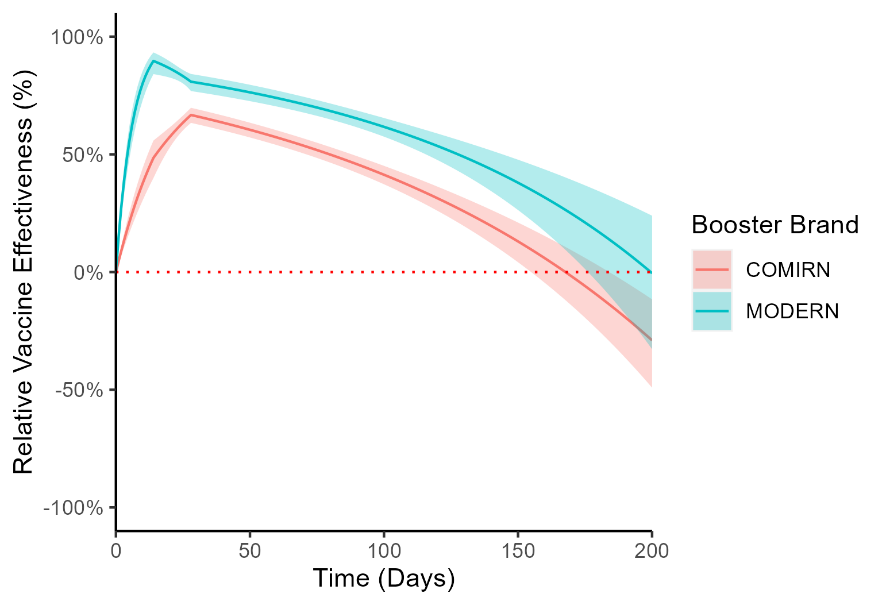 | 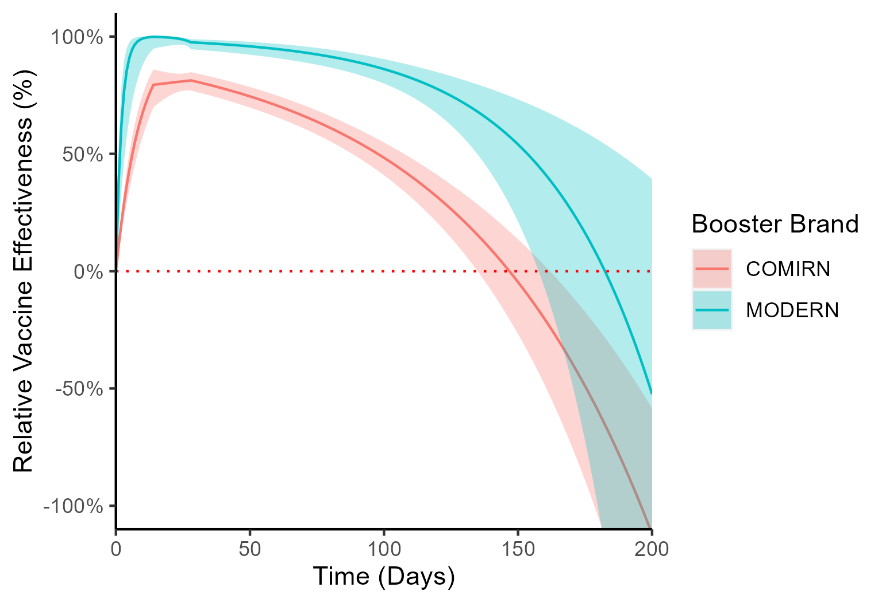 |
| *(c)* | *(d)* |
| 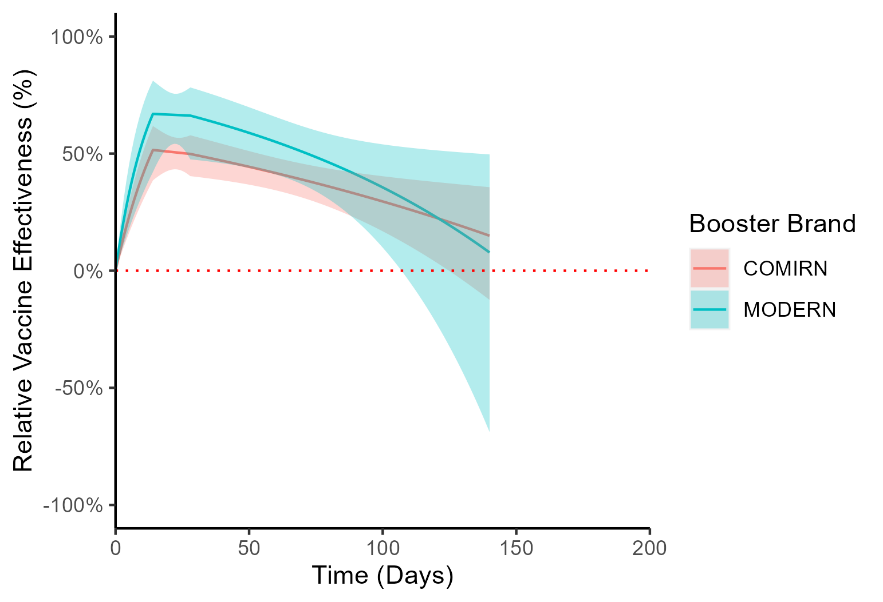 | 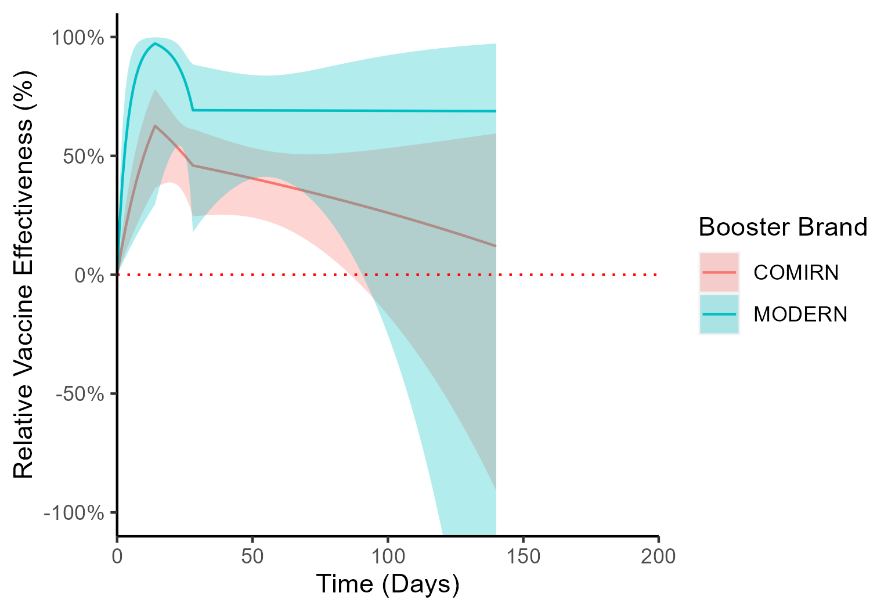 |

Supplementary Figure 1: Relative VE (reduction in hazard) of three vs. two COVID-19 vaccine doses against hospitalisation (a) and death (b) in the Omicron BA.1/2 period, and four vs. three COVID-19 vaccine doses against hospitalisation (c) and death (d) in the Omicron BA.4/5 period, for individuals aged ≥65 years, stratified by brand of booster (estimates in green correspond to Spikevax boosters, estimates in red correspond to Comirnaty boosters). Models are adjusted for age, gender, socio-economic status, Aboriginal and/or Torres Strait Islander identity, whether the individual’s primary vaccination course was with an mRNA or non-mRNA vaccine (for comparisons of three vs. two doses only), and Charlson comorbidity index.

| *(a)* | *(b)* |
| --- | --- |
| 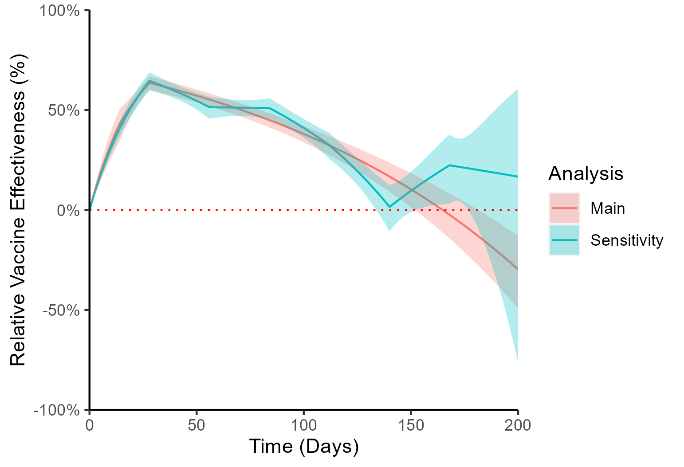 | 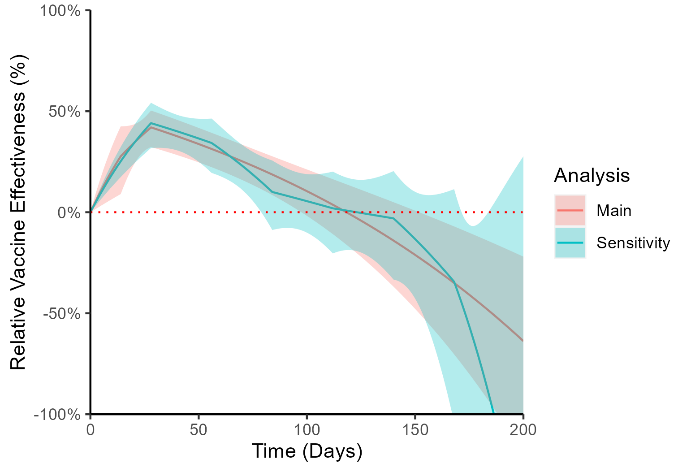 |
| *(c)* | *(d)* |
| 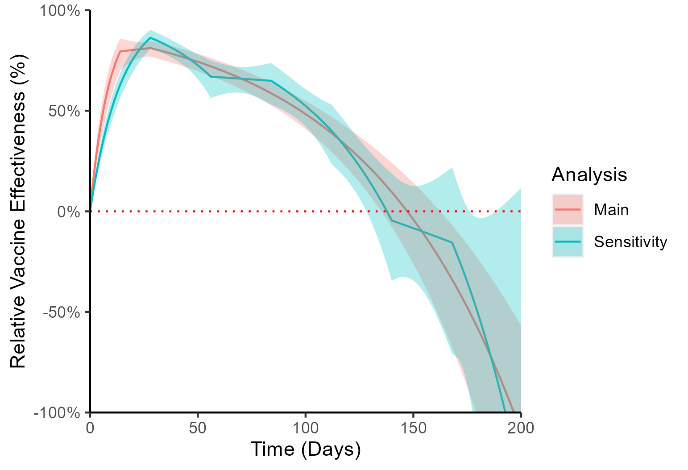 | 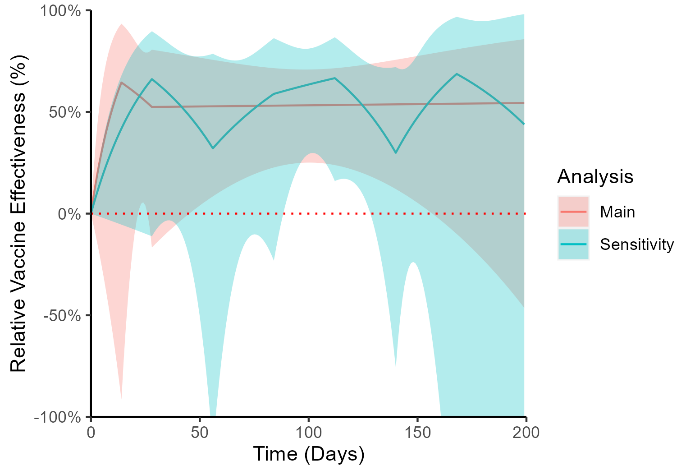 |

Supplementary Figure 2: : Adjusted relative vaccine effectiveness of three vs. two COVID-19 vaccine doses against hospitalisation for individuals aged ≥65 years (a) and 50–64 years (b), and death for individuals aged ≥65 years (c) and 50–64 years (d) from 1 December 2021 to 19 June 2022, with changepoints placed at 4, 8, 12, 16, 20, and 24 weeks (green) compared to the primary analysis (red). Models are adjusted for age, gender, socio-economic status, Aboriginal and/or Torres Strait Islander identity, whether the individual’s primary vaccination course was with an mRNA or non-mRNA vaccine, and Charlson comorbidity index.
